# Stakeholder perspectives on the feasibility and uptake of the cardiovascular polypill in South Africa

**DOI:** 10.1101/2025.10.28.25338994

**Authors:** Trudy D Leong, Bey-Marrie Schmidt, Liesel Zühlke, Robyn Curran, Jane Simmonds, Ntobeko A. B. Ntusi, T mara Kredo, Lara Fairall

**Affiliations:** Health Systems Research Unit, South African Medical Research Council, Parow Valley, South Africa; Division of Health Systems and Public Health, Department of Global Health, Stellenbosch University, Stellenbosch, South Africa; Division of Epidemiology and Biostatistics, Department of Global Health, Faculty of Medicine and Health Sciences, Stellenbosch University, Stellenbosch, South Africa; Office of the President, South African Medical Research Council, Parow Valley, South Africa; Knowledge Translation Unit, Department of Medicine, University of Cape Town, Cape Town, South Africa; Division of Cardiology, Department of Medicine, University of Cape Town, Cape Town, South Africa

**Keywords:** CV polypill, implementation, South Africa, Primary Healthcare, CV prevention, management

## Abstract

**Background:** Cardiovascular disease (CVD) remains the leading global cause of death, disproportionately affecting low- and middle-income countries (LMICs). Complex treatment regimens compromise adherence and health system delivery. Cardiovascular (CV) polypills, combining a statin with two/three antihypertensives, can improve adherence, risk factor control, and outcomes. However, uptake remains low despite inclusion in the 2023 World Health Organization Essential Medicine List. In South Africa, where CVD and multimorbidity are rising, polypills are unavailable in the public sector, with limited use privately. Understanding stakeholder perspectives is essential to guide implementation.

**Objective:** To explore stakeholder perspectives regarding the feasibility, acceptability, and potential uptake of CV polypills in South African primary care.

**Methods:** We conducted semi-structured interviews with 15 purposively sampled stakeholders from clinical, policy, regulatory, industry, and advocacy sectors. Data were thematically analysed using ATLAS.ti, with reflexivity, peer debriefing, and participant validation to ensure rigour.

**Results:** Participants acknowledged polypill’s potential to improve adherence, simplify treatment, and strengthen prevention, but identified key barriers, including formulation challenges, limited incentives, regulatory complexity, inequitable access, and insufficient local evidence. Pharmacological strategies were viewed as complementary to prevention measures such as risk screening and lifestyle modifications. Participants advocated for context-specific research, coordinated policy action, streamlined regulation, equitable procurement, and advocacy-led demand generation, with government leadership deemed essential.

**Conclusion:** CV polypills offer a promising strategy to improve CVD outcomes in LMICs, but impact depends on overcoming systemic barriers and generating local evidence. Coordinated, multisectoral action is critical to achieve scalable and equitable implementation in South Africa and similar settings.

**Contribution:** This study advances understanding of the clinical, behavioural, and system-level factors shaping the adoption of CV polypill in South Africa, providing actionable insights to guide future research, inform policy innovation, and support scalable implementation of cardiovascular prevention strategies across LMIC settings.

## Introduction

Cardiovascular disease (CVD), including myocardial infarction and stroke, remains the leading cause of death worldwide, causing over 17 million deaths annually.^1^ The burden is disproportionately high in low- and middle-income countries (LMICs), which account for more than 80% of premature CVD deaths.^2^ This growing epidemic reflects a complex interplay of social, environmental, commercial, and genetic factors,^3^ posing a significant challenge for achieving Sustainable Development Goals (SDG) 3.4, a one-third reduction of premature non-communicable disease (NCD) mortality by 2030, and universal health coverage (UHC).^4^

In 2001, the World Health Organization (WHO) introduced the concept of the cardiovascular (CV) polypill as a low-cost, scalable, fixed-dose combination (FDC) of multiple cardioprotective agents, to reduce cardiovascular events.^5^ Wald and Law further developed this concept in 2003, proposing a FDC containing aspirin, a statin, three antihypertensive agents, and folic acid, for widespread primary prevention.^6^ Subsequent evidence and evolving clinical guidelines have shifted the polypill paradigm toward secondary prevention, typically combining a statin with two or more antihypertensive agents while excluding aspirin and folic acid. ^7-9^

Randomised controlled trial evidence demonstrates that fixed-dose CV polypills improve treatment adherence, strengthen risk factor control, and reduce major adverse cardiovascular events (MACE) compared with placebo or standard care, across prevention settings. In the PolyIran trial,^10^ n=6838, polypill use resulted in a 34% reduction in MACE compared to lifestyle advice (HR 0.66, 95% CI 0.55 to 0.80), while the TIPS-3 trial,^11^ n=5713, reported a 31% reduction when combined with aspirin compared to placebo (HR 0.69; 95% CI 0.50 to 0.97). In secondary prevention, the SECURE trial,^12^ n=2,499, reported a 24% reduction in composite outcomes of cardiovascular death, non-fatal myocardial infarction, non-fatal stroke or urgent revascularisation, (HR 0.76; 95% CI 0.60 to 0.96) and a 33% reduction CV mortality (HR 0.67; 95% CI 0.47 to 0.96) compared with guideline-directed single-agent usual therapy.

An application to include CV polypills on the WHO Model List of Essential Medicines (EML) was submitted,^13^ with the systematic review evidence later published in 2024.^14^ In 2023, three *“Fixed-dose combinations for prevention of atherosclerotic cardiovascular disease*” were included in the WHO EML: 1) acetylsalicylic acid plus atorvastatin plus ramipril, 2) acetylsalicylic acid plus simvastatin plus ramipril plus atenolol plus hydrochlorothiazide, and 3) atorvastatin plus perindopril plus amlodipine. All components except acetylsalicylic acid were listed as therapeutic alternatives, reflecting the WHO’s emphasis on pharmacological equivalence rather than proprietary products. Despite this policy milestone, translation into practice in LMICs remains limited, constrained by fragmented procurement systems, prescriber inertia, concerns about over-medicalisation, and limited interest from the pharmaceutical industry.^15^ In South Africa, where the incidence of multimorbidity and CVD mortality is on the rise,^16^ only one cardiovascular polypill (Triveram®-atorvastatin/ perindopril/ amlodipine) has been registered since 2021. Currently, its availability is limited to the private sector due to pricing and access barriers.^17^

It is important to differentiate the CV polypill concept from fixed-dose combination antihypertensives, commonly referred to as single-pill combinations (SPCs). SPCs typically consist of two or more antihypertensives but exclude lipid-lowering medications or antiplatelet agents such as aspirin.^8^ While SPCs are commonly prescribed in high-income countries and in South Africa’s private sector, uptake in publicly funded health services in LMICs remains limited, despite inclusion the WHO EML in 2019.^18,19^

This study explores perspectives of key South African stakeholders, including policymakers, regulators, professional associations, pharmaceutical industry representatives, medical scheme funders, and implementation experts, on the feasibility, acceptability, and scalable pathways for introducing CV polypill across public and private sectors to address the growing CVD burden.

## Methods

### Study Approach

This qualitative study explores stakeholder perspectives on the use of the CV polypill in the South African health sector, through a health system and services lens. This approach facilitates an in-depth understanding of stakeholders’ preferences, concerns, and suggestions related to the implementation of the CV polypill.

### Study sample

We purposively sampled 15 interviewees, guided by the principle of data saturation.^20^ The sample included senior policymakers, guideline developers, professional association leaders, healthcare funders, regulatory authority technical experts, academics, clinicians, pharmacists, a nurse, and advocacy representatives – all of whom held multiple roles (Table 1). Targeted stakeholder mapping identified key stakeholders with relevant expertise across the public and private sectors, supplemented with snowball sampling to capture diverse views. As formative research, this sample aimed to capture a wide range of perspectives rather than aiming for exhaustive representation, to inform a prospective implementation study.^21^

**Table 1.** Study participant characteristics.

| ID | Professional training and current roles | Setting |
| --- | --- | --- |
| 1 | Specialist clinician, researcher, policy maker | Public sector |
| 2 | Specialist clinician, researcher, academic | Public sector |
| 3 | Specialist clinician, researcher, academic, regulatory technical specialist | Public & private sectors |
| 4 | Pharmacist, researcher, academic, regulatory technical specialist, policy maker | Public & private sectors |
| 5 | Specialist clinician, researcher, academic, advocacy representative | Public & private sectors |
| 6 | Pharmacist, policymaker | Public sector |
| 7 | Specialist clinician, policy maker | Public sector |
| 8 | Specialist clinician, policy maker | Public sector |
| 9 | Pharmacist, health policy analyst, policy maker | Private sector |
| 10 | Nurse, policy maker | Public sector |
| 11 | Pharmacist, policymaker | Public sector |
| 12 | Pharmacist, regulatory technical specialist, advocacy representative | Public & private sectors |
| 13 | Specialist clinician, researcher, advocacy representative | Public & private sectors |
| 14 | Pharmacist, advocacy representative | Public sector |
| 15 | Pharmacist, advocacy representative | Public sector |

### Data Management

We conducted virtual semi-structured interviews in English between 31 October 2024 and 18 February 2025. Participants received study information and consent forms, and provided written informed consent, including permission to audio record, prior to the interview. Verbal consent was reconfirmed at the interview start. Guided by the semi-structured interview guide, interviews explored the feasibility, acceptability, and potential uptake of the CV polypill in South African primary care. TDL, TK, and BMS conducted the interviews, which ranged from 30 to 60 minutes. Interviews were audio-recorded and securely stored on password-protected SAMRC servers with daily backups. Access was restricted to authorised study personnel to ensure confidentiality.

### Data Analysis

Verbatim transcripts were analysed thematically using ATLAS.ti, version 9.19.1 (ATLAS.ti 2025) by TDL and BMS. An inductive approach guided thematic analysis, aiming to capture participants’ lived experiences and perspectives. Thematic analysis followed Braun and Clarke’s six-phase framework: (1) familiarisation with the data, (2) generating initial codes, (3) searching for themes, (4) reviewing themes, (5) defining and naming themes, and (6) producing the report.^22^ Five participants across private and public sectors, academia, policy, regulatory, and clinical practice were invited to verify the emerging themes, of which three consented. To ensure the trustworthiness of the analysis, we independently coded an initial subset of transcripts and reconciled discrepancies to refine the coding framework.^23^ Reflexivity was embedded throughout the process via regular peer-debriefing, a maintained audit trail, and ongoing reflexive note-taking during data collection and analysis.^24^ Particular attention was paid to the researchers’ positionality and potential influence on interpretation, given the possibility of prior familiarity with some participants. The research team brought a diverse range of professional and disciplinary perspectives, including a policy specialist, social scientist, pharmacologist, and cardiologist. This range of expertise provided both critical reflexivity and complementary skills relevant to data interpretation and contextual understanding.

### Ethical Considerations

The study was approved by the South African Medical Research Human Research Ethics Committee (approval EC011-6/2024) and follows South Africa’s Protection of Personal Information Act^25^ and the Declaration of Helsinki (2024).^26^ Before the study started, permission to interview government officials was received from the South African National Department of Health. Participation was voluntary, with participants free to withdraw at any time without consequences. Confidentiality was upheld through anonymising the transcripts and reporting.

## Results

Of the 21 individuals invited, 15 participated in interviews. Participants held diverse professional roles, including as specialist clinicians (n = 7), pharmacists (n = 7), policymakers (n = 8), researchers (n = 6), academics (n = 4), advocacy representatives (n = 4), regulatory technical specialists (n = 3), a nurse (n = 1), and a health policy analyst (n =1). Many reported experiences across various levels of care in both public and private health sectors. Nine participants worked primarily in the public sector, five in both public and private health sectors, and one exclusively in the private sector. Fourteen were involved in primary healthcare. Pharmaceutical industry and general practitioners were underrepresented, and private-sector perspectives were limited. Participant characteristics are summarised in Table 1.

From our analysis, seven overarching themes emerged:

1. Perspectives and experiences with the CV polypill
2. Awareness of historic challenges to market adoption
3. Views on appropriate clinical indications
4. Perceived benefits and limitations
5. Operational considerations for implementation
6. Advocacy needs and equity considerations
7. Priorities for future research

### Perspectives on the CV polypill

Participants expressed a broad range of views on the value of the CV polypill, but all recognised CVD as a major public health priority. Many supported expanding access, citing improved adherence, reduced pill burden, and potential environmental benefits. As one participant remarked, *“I’ve been an advocate of the polypill for cardiovascular disease for a very long time”* (P12), while another described adherence challenges illustrated by *“bags full of unused medication”* (P8).

Several participants emphasised the importance of affordability and the need *“to tackle patent barriers*… *to accelerate access in low- and middle-income countries”* (P15). The necessity of population-level strategies was also highlighted: *“Unless we do something at a population level, we’re not going to manage health”* (P5). However, several stressed the need for more local evidence and cost-effectiveness data. A few were unfamiliar with the CV polypill, reflecting its limited availability in South Africa. One remarked, *“You mentioned the first polypill registered by SAHPRA…is an innovator product, not a generic”* (P12), raising further concerns about affordability. Participants also highlighted inequities in the availability of CV FDCs (including SPCs for hypertension) between public and private sectors, perceived as a barrier to equitable care

### Historic challenges of bringing the CV polypill to market

Participants attributed the slow uptake of the CV polypill to a combination of scientific, economic, and systemic challenges.

#### Scientific and formulation barriers

Participants consistently reported that early development was complicated by drug-drug interactions, particularly between aspirin and statins; instability of certain antihypertensive agents; and the market withdrawal of a widely used statin over safety concerns. One participant remarked, *“*…*[what is] added to the WHO [Essential Medicines List]…is not a polypill. It doesn’t fit with the original intent*… *It’s merely a fixed-dose combination”* (P4), illustrating how pharmacological complexities reshaped the CV polypill’s composition and clinical objectives.

#### Economic disincentives and market dynamics

The limited commercial value of generic CV polypills was cited as a barrier to pharmaceutical investment. As some noted, *“It might not be economically viable for big pharma to produce a pill that might impact on their market share”* (P8), especially without *“a clear market, a clear route to scale up”* (P13). Limited industry awareness and engagement were also perceived to reinforce a self-perpetuating cycle of low demand and limited supply.

#### Policy, advocacy, and institutional gaps

Policy-wise, participants described fragmented advocacy and unclear institutional mandates: *“To engage industry around our needs… we’re not doing that, and it comes back to the old question of whose role this is… The advocacy component is critical, and it rests with government” (P12)*. Participants perceived the absence of political commitment and strategic leadership as significant as scientific or commercial barriers in shaping the polypill’s trajectory.

### Indications for the use of the CV polypill across the prevention and management spectrum

Participants consistently emphasised that while pharmacological strategies, such as the polypill, hold value, they are insufficient without complementary prevention efforts, such as lifestyle interventions, risk screening and stratification, and robust follow-up systems.

#### Screening and risk stratification for primary prevention

Participants described systematic CVD risk screening as “*very poorly done*” in the public sector with little comprehensive risk assessment (P8). Furthermore, population-level surveillance data was limited, as one participant noted, “*If you ask me how many patients are hypertensive in the country now, I won’t be able to tell you*” (P10). Such diagnostic gaps were identified as major obstacles to achieving national targets and the United Nations Sustainable Development Goals. Participants proposed simple, validated tools, such as point-of-care cholesterol testing and BMI-based risk scoring, as feasible strategies to improve risk identification and guide earlier intervention.

#### Lifestyle interventions

Participants regarded lifestyle interventions, including behavioural counselling and education on diet, physical activity, smoking cessation, and weight management, as essential but challenging to deliver in “*overburdened [facilities]*” (P11). As one participant noted, *“You can’t just take [the CV] pill and expect all the health outcomes as promised”* (P9). While these approaches were seen as empowering patients to take ownership of their health, many acknowledged that social and economic barriers often limit behaviour change. Participants also framed their emphasis on lifestyle approaches within a broader reluctance to medicalise CVD risk, noting low health-seeking behaviour among asymptomatic individuals. To address these challenges, several suggested prioritising high-risk groups, such as those with hypertension, diabetes, or on antiretroviral therapy, as a more feasible strategy to integrate behavioural counselling and risk management into routine care.

#### Secondary prevention and the role of pharmacotherapy

Most participants supported CV polypills for secondary prevention but emphasised the need for locally relevant evidence rather than extrapolation from global trials. As one participant explained, “*there isn’t a simple translation of the WHO model list into South African practice*” (P4). Lessons from the CREOLE trial,^27^ though focused on hypertension, demonstrate that *“the vast majority achieved control on two agents by two months,”*. This was particularly with combinations such as a calcium channel blocker (CCB) and a diuretic (P5), which were suggested to be as more applicable to African contexts.

Parallels with HIV and TB treatment were frequently drawn, where FDCs are now standard practice. One participant remarked, *“Combined medications…for chronic conditions are essentially a must”* (P5). However, there was broad consensus that aspirin use should be limited to secondary prevention as an add-on therapy, noting the adverse effects, associated with aspirin.

Preferred formulations for primary prevention formulations included statin-based combinations (e.g., rosuvastatin or atorvastatin) with two antihypertensives (e.g., ACE-inhibitor/ARB plus CCB, or CCB plus thiazide), while cautioning about potential drug-drug interactions with ARVs. For secondary prevention, patients already stabilised on chronic therapy were seen as ideal candidates, reducing the need for frequent statin titration. Flexibility in dosing and selecting component combinations was viewed as essential to address diverse comorbidity profiles. As one participant explained, *“we need to ensure that we have a range of fixed-dose combination polypills that would target the relevant risk factors”* (P9). Few participants argued that primary prevention may be simpler to implement than secondary prevention, which often requires complex combinations of medicines.

#### Economic considerations

Several participants argued that both primary and secondary prevention with a CV polypill could reduce hospitalisation and long-term healthcare costs: *“…giving them*…*primary prevention [prophylaxis]*…*prevents hospitalisation*…*it is cost effective compared to waiting for the events to happen”* (P14). Others cautioned that formulation choices might be driven more by commercial priorities than patient needs: *“…the drugs that are chosen*…*are what makes commercial sense and not necessarily what’s going to make the best sense for the patient”* (P7). Concerns were also raised about misalignment with existing public-sector treatment guidelines, including the need for robust local cost-effectiveness analyses and deliberate policy integration: *“our standard treatment guidelines don’t necessarily have those specific [medicine] combinations”* (P11).

### Hypertension single-pill combination as a practical entry point

Participants frequently identified SPCs as a practical and strategic entry point for introducing CV FDCs in the public sector context. SPCs were widely perceived to offer advantages for adherence, tolerability, and efficacy by simplifying regimens and enabling dose-sparing approaches.^28,29^ Their effectiveness in African settings was consistently highlighted.^27,30,31^

However, participants observed that SPCs have not generated the same sense of urgency for scale-up seen with HIV or TB FDCs: *“TB/HIV, for example, there is a concern that leaving one drug out of a combination would result in resistance. In hypertension, you don’t have the same externality argument* (P4). Overall, SPCs were seen as “*a no-brainer… there would be an immediate benefit for public health*” (P2), with potential to achieve blood pressure control in “*approximately 80% of patients”* (P13), by shifting practice away from stepwise titration toward simplified fixed-dose regimens. Concerns remained, however, regarding intolerance to individual components and the need for multiple formulations.

### Weighing the pros and cons of using the CV polypill

Participants articulated a nuanced perspective of the CV polypill, recognising its potential to transform prevention and long-term management of CVD while noting clinical, behavioural, economic, and systemic limitations. Table 3 summarises the main advantages and disadvantages of the CV polypill across these domains, linked to participant perspectives with existing evidence. Overall, the CV polypill was viewed as a promising intervention, but its impact was seen to be dependent on appropriate patient selection, alignment with evidence-based guidelines, robust monitoring systems, and coordinated policies and market incentives to facilitate effective scale-up.

### Operational dynamics of introducing CV polypill

Participants described the introduction and scale-up of CV polypills in South Africa as a complex, multi-stage process requiring coordination across regulatory, policy, procurement, and health system domains. Key sequential steps included early stakeholder engagement, regulatory approval, policy adoption, supply chain preparation, and phased implementation with clearly defined roles and responsibilities. Many participants noted that strategies could draw significantly from lessons learned in previous FDC roll-outs.

#### Regulatory approval

Participants consistently identified regulatory approval as a prerequisite for introducing CV polypills and SPCs. As one participant noted, *“a polypill application [should] meet the actual basic requirements, [outlined in] SAHPRA’s guidelines”* (P3). Several highlighted that, once patents expire, generic FDC formulations may qualify for expedited approval based on bioequivalence data if all components are already registered, without the need for further clinical trials. Participants cautioned that the inclusion of a CV polypill on the WHO EML alone was seen as insufficient for market entry, and highlighted the need for proactive leadership, particularly from the National Department of Health (NDoH). Clear product specifications and pricing expectations were viewed as critical to stimulate engagement with manufacturers: *“*… *[if the NDoH requested], ‘We want these kinds of criteria, and by the way, it’s got to cost this at a given scale’, I think you’d have a much more responsive R&D system”*(P13). At the same time, participants pointed out that the low profit margins and risk aversion among generic manufacturers necessitate a clearly defined market pathway.

#### Implementation and access

- *Information gaps* Limited and fragmented information on CV polypills was widely cited as a barrier to implementation. Participants remarked on the absence of patient information leaflets in public sector dispensing, incomplete product data, and challenges in defining public sector tender specifications as factors impeding uptake: “*finding comparable combinations was quite complex… with many questions about quantification and implementation”* (P11). In the private sector, *“formulary listing… guides prescribing”* (P9), and evidence-based marketing were viewed as essential for strengthening prescriber confidence.
- *Prescribing and dispensing practices* Participants further noted that switching patients from multiple individual medicines to an FDC requires a new prescription, as current legislation permits generic but not therapeutic substitution. As one participant explained, *“…regulations at the moment don’t allow pharmacists to substitute* … *molecules within a particular therapeutic area*…*we would need to revisit the regulations”*(12). Several participants also reflected on how remuneration models shape prescribing behaviour. Private sector fee-for-service systems were seen to incentivise personalised prescribing, frequent visits, and multi-pill dispensing to maximise revenue: *“it’s a whole…industry of paying for mediocrity…more fees from separate pills than combination pills… pharmacist co-pays dominate…[and] doctor individualism [drives] personalisation]”* (P13). In contrast, the public sector’s Central Chronic Medicines Dispensing and Distribution (CCMDD) programme, which bills chronic medication per parcel rather than per item, was regarded as more cost-efficient and better aligned with polypill implementation, especially for stable patients: *“it’s cheaper…chronic dispensing is per parcel, not per item…I see huge benefit… but it must be done with care, and it must be done slowly”* (P6).
- *Lessons from other programmes* The rollout of FDCs for HIV and TB was cited as highly relevant, illustrating the value of simplified regimens, nurse-led care models, and robust information systems. As one participant remarked, “Fixed dose combinations… were really a game changer…for the HIV program” (P11). The previously mentioned CCMDD programme and the WHO/PAHO HEARTS^32^ initiative was highlighted as exemplary models. Implemented across more than 30 Latin American countries, HEARTS operates in both public and private sectors, with many countries incorporating FDCs in national EMLs using simplified, standardised treatment algorithms at the primary care level. Participants also highlighted the role of system incentives in facilitating uptake: “South and Central America is probably one of the…best examples…where they’ve switched over to combination therapy and…overcome physician-led restrictions… the incentives were about treating lots of people well, cost-effectively, and [focusing] on the access and scale of therapy” (P11). Despite these advances, participants expressed concerns about the long-term sustainability of NCD programmes in the absence of dedicated global funding.
- *Governance, procurement, and policy alignment* Participants identified guideline endorsement and inclusion on the EML as essential triggers for procurement and clinical uptake. As one participant explained, *“Normative endorsement can trigger a cascade of events… [with uptake and demand driven ultimately as] a consequence of clinical decision[-making]”* (P14). Although PHC STGs generally recommend FDCs, participants noted their absence from public sector tenders, which continues to limit access. The National Essential Medicines List Committee (NEMLC) was widely regarded as pivotal but context-sensitive in its decisions: *“We consider what the WHO says, but it does not influence the decisions that we make… it still goes through our normal governance processes before it gets onto the formulary”* (P11). Views diverged on NEMLC’s remit, with some advocating a formal evaluation of the CV polypill, and others framing the issue as operational: *“…not a NEMLC issue… it’s all operational”* (P11). Participants remarked that although EML inclusion triggers structured procurement, from specification to contracting, uptake remains limited by poor forecasting, scarce utilisation data, clinician acceptance uncertainty, and weak health information systems. The need for stronger market intelligence to inform procurement strategies was mentioned: *“We plan to survey provinces on preferred combinations, distribution strategies, and whether they would pay more for a combination drug… most have very limited resources”* (P11). In the private sector, participants indicated that access to the CV polypill depends on medical scheme formularies, clinical entry criteria, and pricing, all of which influence prescriber choice and patient uptake: *“…formulary status would definitely guide reimbursement…that would be guided by …comparative costing versus the individual agents as well as the clinical entry criteria per condition”* (P9). Participants called for stronger research–policy collaboration but cautioned that direct involvement by the NDoH in manufacturing could create conflicts of interest: *“[It] conflates the responsibilities of purchaser and producer”*(P11). This concern contrasts with the supportive stance of the Department of Trade, Industry, and Science and Innovation, which actively promotes local manufacturing initiatives. Global mechanisms such as the Medicines Patent Pool (MPP) were seen as key enablers of generic access through voluntary licensing and technology transfer. As one participant explained: *“…because [CVS polypills/FDCs] were identified as relevant intervention[s] by the World Health Federation, [and are listed]…on the…WHO EML…[the MPP reviewed the case and determined]…that it was not necessarily a patent intervention, but more trying to understand…from a technology transfer perspective, [if support could be provided to]…expand the manufacturing capacity in low-income countries*” (P15).
- *Coordinated systems for implementation* Participants emphasised practical challenges in implementing CV polypill strategies in primary care required coordinated action *“from clinicians to policymakers”* (P1). Contrary to some views that SPCs offered a simpler entry point, some thought that aspirin–statin polypills would offering the greatest scale-up impact for patients not on full guideline-directed therapy, described as “*…the majority of people in every country”* (P13). Nurse prescribing in South Africa was further seen as *“particularly promising [for expanding access]”*(P13). Public sector tenders, given their scale, were regarded as strategic tools for shaping markets and negotiating pricing. As one participant noted, the national government should leverage this power, emphasising that “*[price] negotiations*… *must happen*… *between the Affordable Medicines Directorate [and the manufacturers]”* (P6).

### Enhancing equity and advocacy for CV polypill implementation

Equity gaps emerged as a recurring concern, with access to SPCs and CV polypills largely confined to the private sector. Participants emphasised that addressing these inequities requires coordinated efforts across clinical guidelines, regulatory processes, procurement systems, and broader health system governance, noting dispensing efficiencies, *“It’s just one dispensing fee instead of three*… *with an FDC”* (P9).

Several stressed that patient advocacy and education are critical to build demand: *“If we don’t have [the] population saying, ‘I want this,’ we may not get it”* (P5). Drawing parallels to HIV treatment scale-up, participants argued that community mobilisation and treatment literacy campaigns could play a similar role in CVD advocacy. As one participant noted, “*People who might need to take 20 tablets a day… might want to fight for having one”* (P1). Participants also highlighted the role of government leadership: *“The minister should be telling the pharma companies*… *they need to be developing these products”* (P12).

In summary, integrated efforts were described as improving medicine access with robust patient education, fostering empowerment, adherence, and sustained demand for the CV polypill.

### Future research priorities

Participants consistently highlighted the need for context-specific research tailored to South Africa’s unique epidemiological profile. They argued that stroke, driven predominantly by hypertension rather than atherosclerosis, remains the most common cardiovascular presentation among individuals of African ancestry: *“We don’t know a lot about vascular disease amongst people of African ancestry… and it seems to be much more related to strokes than to ischaemic heart disease and much more linked to hypertension than to dyslipidaemia, though I’m unsure of the evidence base for statins”* (P1). HIV and obesity were also identified as major compounding risk factors: *“…maybe one could consider the CV polypill for patients at risk — for example, all our patients on chronic ARVs are at risk for cardiovascular disease”* (P7), which could inform research design and intervention strategies.

There was strong consensus that future research must move beyond extrapolating global evidence and instead generate context-specific data on the safety, efficacy, and cost-effectiveness of FDCs, including hypertension SPCs and the CV polypill, in local high-risk populations. Participants saw particular value in studies designed to determine optimal drug combinations, clinical outcomes, adherence impacts, and long-term health system implications. “*I’m not clear about what would be the optimal combination*… *that’s what the research is for*… *looking for the hard outcomes”* (P7). Another participant proposed large-scale, locally led clinical trials comparing CV polypills and SPCs to standard care, ideally measuring patient-important clinical endpoints and analysing South Africa’s unique disease patterns: *“I think all three arms would be helpful [to assess]…the polypill in our context…and make it big enough so that there are hard endpoints”* (P2).

Economic evaluation emerged as another priority, with participants emphasising that improved adherence that could translate into reduced hospitalisations and long-term cost savings, should be demonstrated empirically through cost-effectiveness analyses and budget impact modelling to inform procurement and reimbursement decisions: *“That would be a challenge unless adherence could be translated into something more tangible — in terms of cost savings, reduced downstream expenditure, or improved health outcomes”* (P9).

Implementation science research was also deemed critical, particularly to explore strategies for integrating polypills into existing care pathways, strengthening risk stratification approaches, procurement processes, and aligning interventions with standard treatment guidelines: *“How do we stratify risk and build that into our standard treatment guidelines?”* (P4). Some questioned whether CV polypill research currently aligns with national priorities: *“Does [the CV polypill] actually fit our research agenda?”* (P4). Additionally, behavioural and social science research was considered vital to inform patient education, community engagement, and strategies to strengthen health-seeking behaviours: *“Members of the community must take responsibility for their health*… *and to do that, they need to be empowered and capacitated”* (P10).

Overall, participants articulated cautious optimism regarding the potential of CV polypill and SPCs to transform CVD management in South Africa. They highlighted both their potential to improve adherence and outcomes, and the structural, economic, and governance challenges that could limit impact. They emphasised that research priorities should span the clinical, economic, behavioural, and policy domains, and be designed not only to generate evidence but also to inform implementation and scale-up. These insights provide a roadmap for the next phase of research that is contextually grounded, policy-relevant, and directly aligned with health system needs, laying the foundation for a robust evidence base to guide future decision-making.

## Discussion

This study explored stakeholder perspectives on the implementation of CV polypills in South Africa, revealing both considerable promise and persistent challenges. Participants viewed these interventions as valuable for improving adherence, reducing pill burden, and strengthening population-level prevention. However, they highlighted several constraints, including scientific uncertainty, regulatory complexity, limited commercial incentives, fragmented policy leadership, and inequitable access. There was broad agreement that pharmacological interventions alone are unlikely to achieve meaningful population impact without complementary prevention measures such as lifestyle modification, risk screening, and sustained follow-up care.

Evidence from randomised trials, PolyIran,^10^ TIPS-3,^11^ and SECURE^12^ suggests that polypill strategies can improve adherence and reduce MACE and mortality. However, these trials were largely conducted outside SSA, and locally generated evidence remains sparse. African studies, such as CREOLE,^27^ have demonstrated the benefits of hypertension-specific SPCs but have not evaluated statin-containing regimens. Our findings are consistent with implementation research from other LMICs,^8,33^ which identifies similar barriers, including regulatory complexity, fragmented procurement, limited manufacturer engagement, and clinician hesitancy - often more limiting than the pharmacological evidence itself. This study builds on that evidence by situating the CV polypill debate within South Africa’s context, where unique epidemiological patterns, resource constraints, and systemic challenges shape policy and implementation decisions. Notably, participants highlighted that prioritising antihypertensive SPCs could serve as a strategic entry point for broader adoption of CV FDCs in the public sector.

Consensus emerged on the need for locally generated data to inform scale-up, but participants diverged on key strategic questions. Some supported broad use of CV polypill for primary prevention, especially among high-risk groups, while others advocated for a more cautious approach, restricting initial use to secondary prevention among stable patients until robust, context-specific evidence of safety, effectiveness, feasibility, acceptability, and cost-effectiveness becomes available. These differences reflected concerns about aspirin safety in low-risk populations, the need for improved risk stratification, and alignment with treatment guidelines. Views also varied on implementation pathways, with some favouring large-scale local trials prior to adoption and others supporting phased introduction with iterative evaluation. Across all perspectives, there was strong agreement that government leadership is critical to advance regulatory reforms, coordinate policy, shape markets, and mobilise advocacy.

The recently published World Heart Federation (WHF) Roadmap on SPC Therapies^34^ echoes many of these findings, identifying structural barriers, such as manufacturing capacity, affordability, and inconsistent guideline inclusion; and highlighting that evidence alone is insufficient without policy alignment, market shaping, and system readiness. Its practical recommendations, such as regulatory harmonisation, pooled procurement, and clinician engagement, align closely with stakeholder priorities identified in this study.

Importantly, our findings highlight contextual complexities that challenge “one-size-fits-all” implementation approaches. The dual burden of hypertension and HIV, the predominance of stroke over ischaemic heart disease, and stark public-private sector disparities create a health system landscape in which global models may have limited applicability without careful local adaptation. In this context, our study enriches the WHF Roadmap’s global recommendations with granular, ground-level insight and a cautionary perspective on the conditions required for effective implementation in our resource-constrained setting.

### Strengths and limitations

This study provides novel insights into the policy, implementation, and research landscape for CV polypills and SPCs in South Africa, drawing on diverse perspectives from clinical, regulatory, policy, and advocacy domains. Its qualitative design enabled an in-depth exploration of contextual factors shaping adoption and scale-up, many of which are underrepresented in existing literature. However, the study has several limitations. Participants were primarily drawn from national and provincial levels, wand the perspectives of frontline primary care providers may not be fully captured. Industry and private sector perspectives are also limited. Findings are context-specific and may not be directly generalisable to other health systems, although many lessons are transferable. Finally, as with all qualitative studies, the results reflect participants’ perceptions and experiences rather than measurable outcomes. Despite these limitations, the study offers valuable evidence to inform future policy, implementation strategies, and research priorities for CV polypill integration.

### Implications for policy and practice

Our findings highlight that the implementation of CV polypills and SPCs will require coordinated action across multiple levels of the health system. Policy leadership is particularly critical: inclusion of specific CV polypills on the national STGs and EML, and integration into procurement and reimbursement frameworks could enable widespread adoption. Establishing streamlined regulatory pathways, including expedited approval processes for generic formulations based on bioequivalence data, and transparent pricing expectations, may help stimulate market entry and encourage manufacturer engagement. Similarly, strategic use of public procurement mechanisms, such as national tenders and pooled purchasing, could further shape markets, secure affordable pricing, and ensure reliable supply. However, given the anticipated size of the NCD population, sufficient manufacturing capacity will be essential to ensure reliable supply and sustain adherence to long-term pharmacotherapy.

At the service delivery level, integrating polypill strategies within existing chronic disease and primary care platforms has the potential to improve continuity of care and optimise resource use. Lessons from HIV and TB programmes, particularly the value of simplified treatment protocols, nurse-led models, and robust supply chains, offer valuable blueprints for integration of polypills into cardiovascular care pathways.

Sustained advocacy and meaningful patient engagement are likely to be essential in generating demand, shaping policy agendas, and incentivising industry involvement.

Finally, deliberative policies, such as targeted procurement, high-risk population prioritisation through standardised protocols, will be essential to address equity gaps, particularly across public and private sectors, and ensure that the benefits of simplified, evidence-based cardiovascular prevention strategies reach those most in need.

### Research priorities and knowledge gaps

Participants identified several critical evidence gaps that warrant attention to inform effective implementation. These include:

- Clinical effectiveness: Determining clinical effectiveness (including blood pressure control, MACE and mortality), the optimal drug combinations, evaluating safety and adherence, and assessing long-term outcomes in local populations – relative to the standard of care.
- Burden of disease: South Africa’s prevalence of stroke associated with hypertension.
- Implementation science: Identifying strategies for risk stratification, patient selection, procurement and supply chain logistics, and integration into existing care pathways.
- Economic evaluation: Conducting cost–benefit analyses and budget impact assessments to inform policy and reimbursement decisions.
- Behavioural and social science: Designing healthcare worker and patient education and engagement strategies to strengthen adherence and stimulate demand.

Collectively, these research priorities can generate the robust evidence base necessary to support policy and clinical decision-making, ensuring that CV FDCs achieve meaningful and sustained population-level impact.

## Conclusion

This study illustrates the complex clinical, behavioural, and systemic factors influencing the feasibility of CV polypill implementation. Participants acknowledged its potential to improve adherence, streamline services, and expand access, especially in resource-limited settings, while highlighting barriers such as limited dose flexibility, difficulty attributing side effects, clinician resistance, and supply constraints.

Realising this potential will require supportive policies, alignment with national guidelines, and sustained industry engagement to ensure a reliable supply. The polypill should be positioned as a complementary intervention within broader CV prevention strategies, guided by simple, standardised treatment protocols that enable scale-up. Additionally, the availability of multiple fixed-dose combinations with varying strengths will be essential to support individual treatment needs.

Coordinated action across policy, regulatory, procurement, and service delivery domains, underpinned by advocacy, local evidence generation, and community engagement, will be critical for integrating polypill strategies into South Africa’s health systems and advancing equitable cardiovascular disease prevention.

## Ethical considerations

The Human Research Ethics Committee of the South African Medical Research Council granted ethical approval for this study (approval EC011-6/2024).

## Acknowledgements

The authors extend their sincere gratitude to the participants, whose time, insights, and experiences were invaluable to this research and fundamental to the study’s findings.

## Author contributions

Conceptualisation: LF, NN, LZ; protocol development: TK, TDL, BMS, LF, JS, LZ, LF. Data curation: TDL, BS, TK; Analysis and interpretation of the data: TDL, BS; Drafting the initial article: TDL, BS; Revising the article for important intellectual context: All; Review and edit the final draft manuscript: TDL. Final approval of the version of the article to be published: All.

## Data availability

Data supporting the findings of this study are available from the corresponding author, TDL, upon reasonable request.

## Disclaimer

The views and opinions expressed in this article are those of the authors and are the product of professional research. They do not necessarily reflect the official policy or position of any affiliated institution, funder, agency, or that of the publisher. The authors are responsible for this article’s results, findings, and content.

